# EPIDEMIOLOGY OF LEVODOPA-INDUCED DYSKINESIA: PREVALENCE AND ASSOCIATED CLINICAL FACTORS IN LATIN AMERICA

**DOI:** 10.1101/2025.08.25.25334104

**Authors:** Henry Mauricio Chaparro-Solano, Daniel Teixeira-dos-Santos, Emily Waldo, Thiago P. Leal, Miguel Inca-Martinez, Sarael Alcauter, Alejandra Medina-Rivera, Alejandra E. Ruiz-Contreras, Mario Cornejo-Olivas, Koni Mejia-Rojas, Cintia Armas, Pedro Chana-Cuevas, Natalia Rojas, Jorge L. Orozco, Beatriz Munoz Ospina, David Aguillon, Omar Buritica, Sonia Moreno Masmela, Vitor Tumas, Artur F. S. Schuh, Carlos Roberto Rieder, Bruno Lopes Santos-Lobato, Juliana S. Duarte, Susana L. Peña, Mayela Rodríguez-Violante, Ana Jimena Hernández-Medrano, Emilia M. Gatto, Gustavo Andres Da Prat De Magalhaes, Gonzalo Arboleda, Marcelo Andres Kauffman, Sergio Alejandro Rodriguez-Quiroga, Angel Vinuela, Alan Osvaldo Espinal Martinez, Pedro Braga-Neto, Sarah Camargos, Elias Fernandez-Toledo, Cesar Luis Avila, Francisco Eduardo Cardoso, Patricio Olguin, Vanderci Borges, María Valentina Müller, Marcelo Merello, Ana Rosso, Daniel Martinez-Ramirez, Dario Sergio Adamec, Marcela Susana Tela, Grace Helena Letro, Denise H. Nicaretta, Ignacio F. Mata, the Latin American Research consortium on the Genetics of Parkinson’s Disease (LARGE-PD)

## Abstract

**Background:** Although levodopa remains the gold standard treatment for Parkinson’s disease (PD), its chronic use is associated with levodopa-induced dyskinesia (LID), a motor complication that impacts prognosis, quality of life, and treatment costs. Most known LID-associated factors have been identified in European-descendant populations.

**Objectives:** To describe the epidemiology of LID in Latin American and Caribbean (LATAM) countries and assess the relevance of known and novel LID-associated factors in this population.

**Methods:** We conducted a cross-sectional study using data from the Latin American Research consortium on the Genetics of Parkinson’s Disease (LARGE-PD). We included PD patients with information on LID status and levodopa use from eight LATAM countries. LID prevalence was calculated overall and by country. Countries were compared on demographic and clinical variables. Logistic regression was used to identify associations with LID.

**Results:** A total of 3,695 PD patients (58.8% male) were included. Overall LID prevalence was 25.4% [95% CI: 24.06–26.87], ranging from 9.3% in Colombia to 45.1% in Puerto Rico. Prevalence increased progressively with longer disease duration. Country comparisons showed that not all known LID-associated factors explained prevalence differences. In logistic regression, fast disease progression was significantly associated with LID (OR: 1.55, 95%CI: 1.16–2.07), while sex was not (OR: 1.02, 95%CI: 0.87–1.18).

**Conclusions:** This is the largest study on LID epidemiology in LATAM. While some known risk factors remain relevant, others, like sex, do not, underscoring the need for population-specific studies. Future work should integrate environmental, clinical, and genetic data to better understand LID mechanisms.

## INTRODUCTION

Treatments for Parkinson’s disease (PD) are primarily symptomatic, as no disease-modifying therapies are currently available. Among these, levodopa, administered in combination with dopa decarboxylase inhibitors, is considered the gold standard and a first-line treatment for PD [1]. Levodopa is effective in controlling motor symptoms and improving survival [2,3]. However, over time, its use is associated with the development of levodopa-induced dyskinesia (LID), an unpredictable motor complication that can range from mild to severe [1,2,4].

LID is characterized by involuntary, discontinuous and arrhythmic movements that may affect various body regions. It is commonly classified into four main subtypes based on the timing of onset relative to levodopa administration (ON and OFF phases), anatomical distribution and movement patterns [5].

Regardless of type, LID can significantly impact the quality of life of people with PD and their families by increasing the risk of disability, balance impairments, and falls, which in turn raise treatment costs [1,6–8]. Although certain advanced therapies, such as deep-brain stimulation (DBS) and infusion therapies, have proven effectiveness in alleviating LID, access to these interventions is limited, with only a minority of patients able to benefit from them [8]. This is particularly relevant in developing countries, where healthcare services are restricted to economic and technological constraints [9].

The prevalence of dyskinesia has been primarily reported in European populations, with more recent data emerging from Asian countries such as South Korea. Specifically, reported LID prevalence for European countries ranges from 14% to 30% [10–14], whereas in Asian countries it is approximately 40% [15]. However, LID prevalence has been shown to vary widely depending on the duration of PD, with rates gradually increasing over time for both populations [13–15].

In addition to disease duration, other clinical factors have been associated with LID occurrence, including age at PD onset, disease severity, sex, body weight and the daily dose of levodopa therapy [7,14,16]. However, as these findings are mainly derived from non-Latino populations, their relevance to Latino individuals remains unclear. This limits their applicability in clinical decision-making aimed at preventing or delaying the onset of LID.

Moreover, the limited literature on LID in Latino populations primarily comes from studies in Brazil [9,17] and Mexico [18], which often feature small sample sizes, thereby restricting the generalizability of their findings to other Latin American countries.

This study aims to describe, for the first time, the epidemiology of LID across multiple Latin American and Caribbean countries. Additionally, we seek to assess the relevance of previously known LID-associated factors in this population and to explore other clinical variables that may influence the likelihood of developing LID. Understanding these factors is essential for optimizing therapeutic decisions and informing health policies across the region.

## METHODS

We conducted a cross-sectional study using data from the Latin American Research consortium on the Genetics of PD (LARGE-PD) [19]. A total of 4,069 participants with a PD diagnosis confirmed by a movement disorders specialist were recruited between April 2019 and April 2025 across Argentina, Brazil, Chile, Colombia, El Salvador, Mexico, Peru, and Puerto Rico. Only participants with available data on LID status (yes vs. no) and a documented history of levodopa use were included in the analysis. Data corresponds to the April 2025 extraction from the ongoing LARGE-PD study.

Demographic and clinical data were extracted from the LARGE-PD questionnaire administered at the time of recruitment. LID status was primarily based on consistent self-report by participants and, in some cases, confirmed by a movement disorders specialist through observation of dyskinetic movements during the study visit.

Variables relevant to LID were selected for analysis upon previous studies, and include participants’ age, age at onset (AAO), sex, presence of tremor as a presenting motor symptom, dopaminergic treatment dosage and duration, as well as anxiety, a non-motor symptom also previously associated with increased LID risk. The full list of variables is presented in Table 1.

**Table 1.**
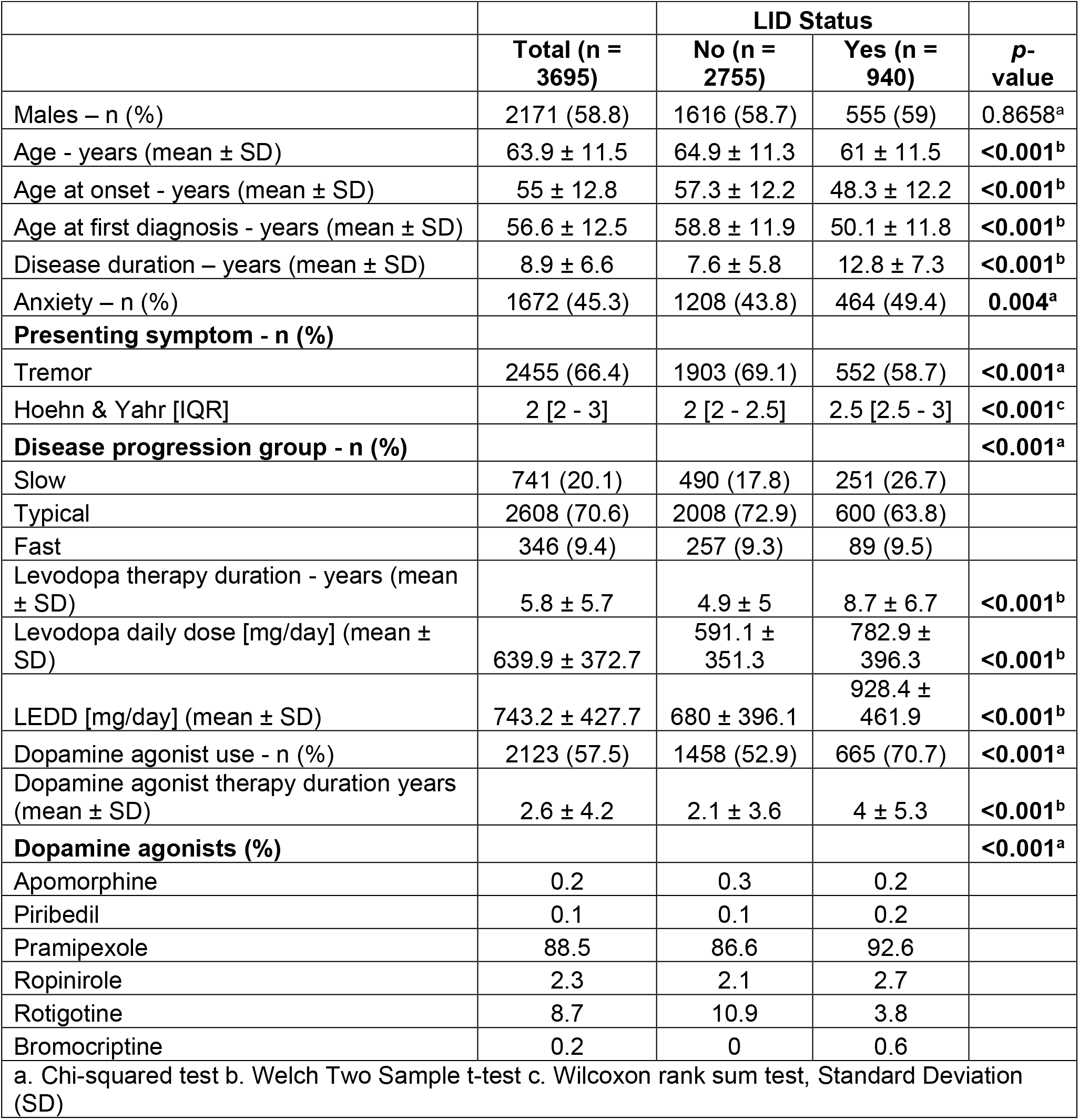
Characteristics of participants with and without LID.

Participants with ≥20% missing data across variables were excluded. Simple imputation was applied for variables with <1% missingness, using the median for quantitative variables and the mode for qualitative variables. Variables with ≥1% but < 20% missingness were imputed using the Multivariate Imputation by Chained Equations (MICE) algorithm [20]. Sensitivity analyses were conducted using the dataset before imputation to ensure robustness of results.

Participants were categorized into two groups based on LID status (yes/no). Disease duration was defined as the number of years from the onset of the first motor symptom to the time of recruitment. For analysis, participants were categorized into five disease duration groups: 0-5 years, 6-10 years, 11-15 years, 16-20 years, or >20 years, following the classification used by Chang et al. [15].

LID prevalence was calculated for the entire cohort and separately by country, stratified by disease duration groups. Prevalence estimates were accompanied by 95% confidence intervals. Additionally, prevalence was also analyzed by sex and AAO (<50 vs. ≥50 years old), following the cut-off for early-onset PD recommended by the Movement Disorders Society (MDS) [21].

Disease progression was defined as a categorical variable using disease duration and Hoehn and Yahr (H&Y) stage at the time of recruitment. Based on the median time to reach H&Y stage 3 reported by Zhao et al. (~8.9 years from stage 1), participants were classified as having “fast” progression if they reached H&Y ≥3 in less than 9 years, “slow” progression if they remained below stage 3 after 9 or more years, and “typical” otherwise [22].

The Levodopa Equivalent Daily Dose (LEDD) was calculated using the most recent formula proposed by Jost et al. [23]. As the LARGE-PD questionnaire only recorded daily doses of levodopa and dopamine agonists, LEDD estimates were based solely on these medications.

Numerical variables were summarized using means or medians, and categorical variables were reported as percentages. Differences between LID status groups were assessed using Chi-square tests for categorical variables and Wilcoxon or t-tests for numerical variables, as appropriate.

Differences in overall estimated LID prevalence between countries, as well as prevalence stratified by disease duration, were evaluated using pairwise Chi-square tests with Bonferroni adjustment. Similarly, average LEDD values, disease duration and AAO across countries were compared using Kruskal-Wallis test, followed by post hoc pairwise comparisons with Dunn’s test and Bonferroni adjustment.

Simple logistic regression analyses were conducted to identify variables associated with a higher probability of developing LID. Variables that reached statistical significance were included in a multiple logistic regression model. Collinearity was assessed before model inclusion. Country of residence was included as a covariate to adjust for potential confounding due to between-country differences. The country with a LID prevalence closest to the cohort-wide average was selected as the reference group.

A significance threshold of p < 0.05 was used for all statistical tests. All analyses were completed using R version 4.3.1.

This study was approved by the Institutional Review Board of the Cleveland Clinic Foundation (IRB # 25-081). In addition, each participating LARGE-PD site obtained approval from its local IRB in accordance with international ethical standards, and all participants provided written informed consent prior to enrollment.

## RESULTS

### Demographics

A total of 3,695 patients with PD (mean age 63.9 ± 11.5 years, 58.8% males) were included in this study. The average disease duration was 8.9 ± 6.6 years (Table 1).

The overall prevalence of LID in the entire group was 25.4%. When analyzed by country, Colombia had the lowest prevalence (9.3%) while Puerto Rico had the highest (45.1%) (Table 2).

**Table 2.** Prevalence of LID among participants overall and by country.

| **Table 2. Prevalence of LID among participants overall and by country** |  |  |
| --- | --- | --- |
| **Country of recruitment** | **N** | **Prevalence (%) [95% CI]** |
| Argentina | 349 | 27.5 [22.82–32.19] |
| Brazil | 759 | 42.6 [39.04–46.07] |
| Chile | 509 | 27.9 [24.00 - 31.79] |
| Colombia | 680 | 9.3 [7.09 - 11.44] |
| El Salvador | 126 | 31.8 [23.62 - 39.87] |
| Mexico | 674 | 18.0 [15.06 - 20.85] |
| Peru | 507 | 22.5 [18.85 - 26.12] |
| Puerto Rico | 91 | 45.1 [34.83 - 55.28] |
| **Overall** | **3695** | **25.4 [24.06 - 26.87]** |

Statistically significant differences in overall LID prevalence were found between most countries, based on pairwise Chi-square tests with Bonferroni adjustment. Colombia was the only country with a significant lower prevalence compared to all others (Supplementary Figure 1). While differences in demographic and clinical characteristics were observed between some countries, these were not consistently present in all comparisons involving those with significantly different LID prevalence (Supplementary Figure 2).

### LID-associated factors

#### Univariate model

Thirteen different factors were found to be significantly associated with LID in the univariate logistic regression analysis, including AAO, disease duration, LEDD, use of dopamine agonists, presence of tremor as an initial motor symptom, history of anxiety or panic as non-motor symptoms, and disease progression rate, among others. Interestingly, sex was not significantly associated with LID (Table 4).

#### Disease duration

A significant gradual increase in LID prevalence was observed across the defined disease duration groups (*p* < 0.05), with the most notable occurring between the first three groups (0-5 years; 6-10 years; 11-15 years). Pairwise comparisons using Chi-square tests with Bonferroni adjustment confirmed significant differences between adjacent groups up to the 16-20 years group, with a non-significant difference between the 16-20 and >20 years groups (Table 3).

**Table 3.** LID prevalence by disease duration groups.

| **Table 3. LID prevalence by disease duration groups** |  |  |  |
| --- | --- | --- | --- |
| **Disease duration group** | **N** | **Prevalence (%) [95% CI]** | **p-value vs. previous group** |
| 0-5 years | 1356 | 8.8 [7.27 – 10.28] |  |
| 6-10 years | 1167 | 25.1 [22.62 – 27.60] | **<0.001** |
| 11-15 years | 675 | 39.0 [35.28 – 42.64] | **<0.001** |
| 16-20 years | 277 | 52 [46.10 – 57.87] | **0.003** |
| > 20 years | 220 | 55 [48.43 – 61.57] | 1 |

LID prevalence by disease duration group was also analyzed by country. This pattern was consistent across most countries, except for Peru and Puerto Rico (Figure 1).

**Figure 1.**
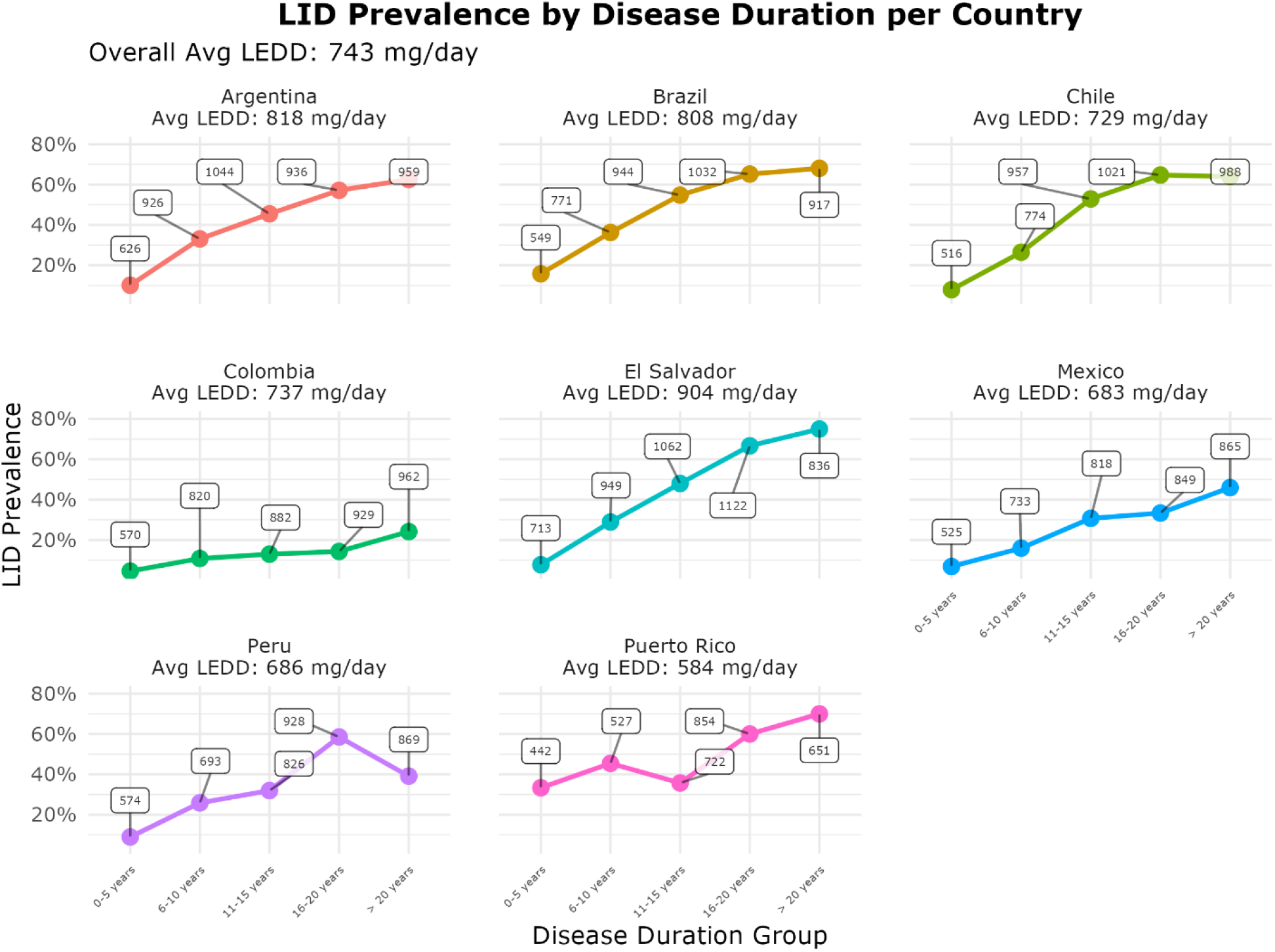
Levodopa-induced dyskinesia (LID) prevalence by disease duration group and country, and average LEDD by country and by disease duration. Each panel shows the proportion of patients with LID (Y-axis) by duration of Parkinson’s disease in years (X-axis) for countries participating in the study. Colored lines represent trends within each country, with points indicating LID prevalence in each disease duration group. White boxes show the average Levodopa Equivalent Daily Dose (Avg LEDD) per disease duration group, and the overall Avg LEDD for each country is shown in the panel title.

#### Age at onset

A similar analysis was conducted by AAO category (<50 vs. ≥50 years old). The overall prevalence of LID was 42.8% among participants with AAO <50 years, compared to 17.4% in those with onset ≥50 years. When stratified by disease duration, LID prevalence remained significantly higher in the younger-onset group across all categories (*p* < 0.001), except in the >20 years group, where the difference was not statistically significant (56.2% vs. 50.0%, *p* = 0.565; Supplementary Table 1).

In both AAO groups, LID prevalence increased with disease duration. Among participants with onset before age 50, prevalence rose from 16.8% in the 0–5 years group to 61.8% in the 16–20 years of disease duration group, followed by a slight, non-significant decline to 56.2% in those with > 20 years (Chi-square test, *p* = 1.00; Supplementary Figure 3). In contrast, among individuals with onset at or after age 50, LID prevalence gradually increased from 7.0% in the 0-5 years group to 50% in the >20-year group (Figure 2).

**Figure 2.**
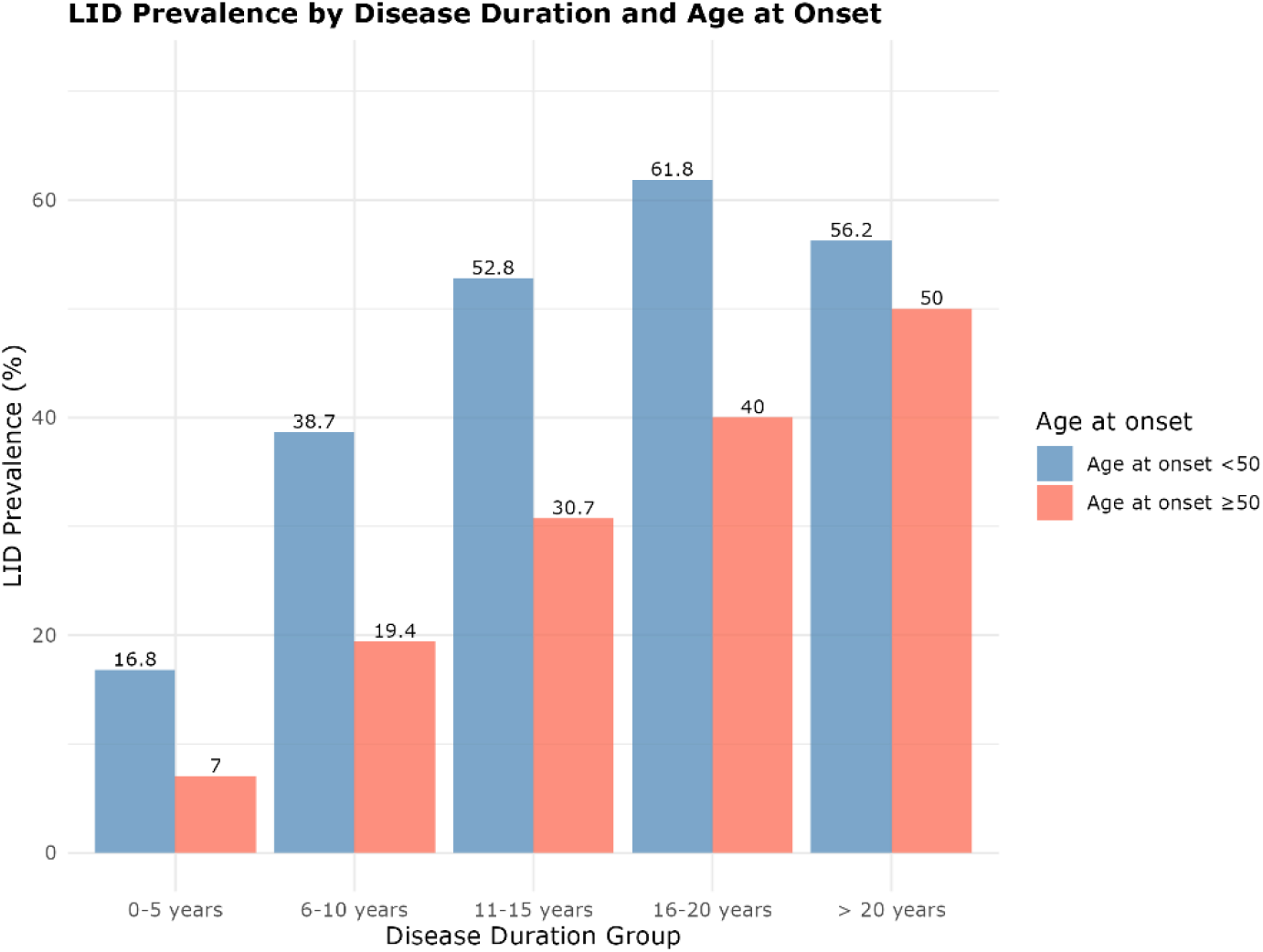
Levodopa-induced dyskinesia (LID) prevalence by disease duration and age at onset. Prevalence of LID (Y-axis) across different Parkinson’s disease duration groups (X-axis), stratified by age at disease onset (<50 vs. ≥50 years-old). Blue bars, age at onset <50 years-old, red bars, age at onset ≥50 years-old.

#### Levodopa Equivalent Daily Dose (LEDD)

While Kruskal-Wallis test indicated a statistically significant difference in average LEDD across countries (*p* < 0.05), not all pairwise comparisons were significant. For example, average LEDD in Colombia was comparable to that in Brazil, Chile, Mexico, or Peru (adjusted *p* > 0.05; Supplementary Figure 2).

#### Multivariable model

A multiple logistic regression model, including all variables independently associated with LID and accounting for collinearity, identified several clinical factors that remained significantly associated after adjustment. Increased odds of LID were observed in participants with younger AAO, a history of anxiety or panic as non-motor symptoms, longer duration of levodopa therapy, higher LEDD, and faster disease progression, whereas tremor as a presenting symptom was associated with decreased odds (Table 4).

**Table 4.**
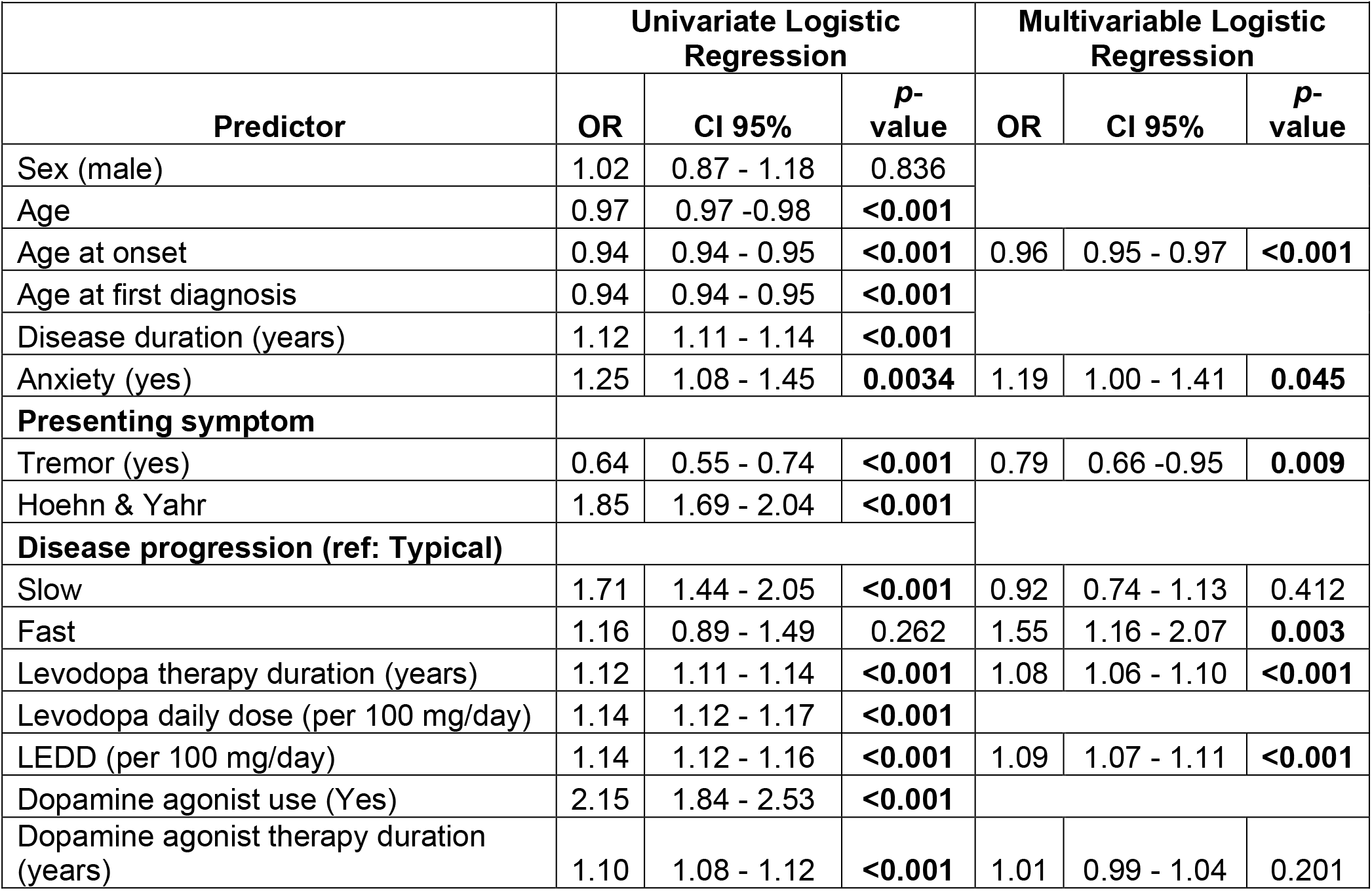
Univariate and multivariable logistic regression analyses of factors associated with LID.

## DISCUSSION

To our knowledge, this is the largest study to analyze the epidemiology of LID in Latin American and Caribbean countries. We estimated an overall LID prevalence of 25.4%, ranging from 9.3% to 45.1% across participating sites, with Colombia and Mexico presenting the lowest, and Brazil and Puerto Rico the highest (Table 2).

Our overall LID prevalence of 25.4% was lower than estimates reported in other populations within and outside Latin America. For instance, Gandhi et al, in a prospective cohort of 3,343 European-descendant participants, reported a LID frequency of 42.1% among those with 8-10 years of disease duration [13]. Notably, their study did not report an overall prevalence, as all values were stratified by disease duration. We therefore used the 8-10 years group for comparison, as it was closest to the mean disease duration in our cohort (8.9 years).

In a smaller retrospective sample of 279 patients in the United States, Turcano et al., found a LID prevalence of 30.1% [10], while Chang et al. reported 43% in a larger sample of 2,571 individuals from South Korea [15]. We did not identify additional previous reports in larger European samples that did not account for AAO or disease duration when reporting LID frequency.

When comparing prevalence by country, our estimate for Brazil (42.6%) fell between two previous reports, 34.7% and 54.9%, from independent cohorts of 187 and 224 participants, respectively. These cohorts corresponded to two non-overlapping sub-samples of LARGE-PD, in which LID was defined as a score of ≥1 on item 4.1 of UPDRS Part IV [9]. Despite differences in LID assessment methods, the similar prevalence estimates suggest good reliability of our combined self-reported and clinician-observed data.

A similar trend of lower LID prevalence estimates was observed when stratifying by disease duration. Although prevalence in the 0-5 years group was comparable between our cohort (8.8%) and that of Chang et al. (8.2%), differences became more pronounced in longer-duration groups. From 6-10 years onward, their reported prevalence nearly doubled ours: 40.7% vs. 25.1% for 6-10 years; 66% vs. 39% for 11-15 years; 74.6% vs. 52% for 16-20 years; and 83.2% vs. 55% for >20 years of disease duration [15].

When examining country-specific differences in clinical variables commonly associated with LID, such as disease duration and LEDD, we found statistically significant differences between some countries, but not all. For instance, Colombia, which had the lowest prevalence, did not differ significantly in disease duration from any country except Brazil, which had the second-highest prevalence. Conversely, when comparing Colombia’s LEDD with the other countries, significant differences were observed with Argentina, El Salvador and Puerto Rico (Table 2; Supplementary Figure 2).

These findings raise the possibility that other factors beyond the well-known clinical variables associated with LID may influence its development differently across countries. Potential contributors could include population differences in genetic variations related to dopaminergic pathways [24], timely access to healthcare [25], and variability in medication availability or prescription practices, particularly the use of immediate-release versus controlled- or extended-release of levodopa formulations. This distinction is important, as non-immediate-release formulations may reduce the pulsatile stimulation associated with immediate-release levodopa, which has been implicated in the pathophysiology of LID [26].

Clinical factors previously associated with an increased odd of developing dyskinesia, such as earlier age at disease onset, longer disease and levodopa treatment duration, and higher LEDD, were confirmed in our cohort. Tremor as a presenting symptom was found to be negatively associated with LID, consistent with prior reports [7,14,17]. In addition, fast disease progression emerged as a potential novel factor associated with the development of LID.

From a biological standpoint, motor complications are primarily driven by the progressive loss of dopaminergic neurons, which diminishes the brain’s capacity to buffer dopamine. This is facilitated by an increased dopamine availability, which is processed by serotonergic neurons [26]. This mechanism may help explain our observation that patients with faster disease progression, who experience these neurobiological changes more rapidly, develop LID more frequently than those with slower progression.

The association observed between AAO and LID in our analysis adds further evidence to the well-established link between the younger onset of PD and increased risk of LID. This may reflect a combination of treatment-related and intrinsic predisposition. Although early-onset PD is generally associated with slower motor progression, our findings, along with previous reports [15], suggest that these patients remain at high risk for LID due to factors other than the accelerated neurodegeneration typically linked to faster disease progression [26].

One possible explanation is the longer disease duration, often accompanied by an early and rapid increase in dopaminergic dosage, likely driven by the need to control motor symptoms and maintain daily functioning, including occupational and family responsibilities [27]. In addition, genetic susceptibility to the development of LID may also be relevant. Previous evidence suggests that PD patients in the highest quartile of Polygenic Risk Scores (PRS), which aggregate PD-associated genetic variants, have a higher risk of developing LID [24]. Moreover, a separate study has reported that higher PD-PRS is associated with earlier onset of LID [13].

Our findings propose that faster disease progression and younger AAO represent complementary risk profiles in Latin American populations, each independently increasing susceptibility to LID through distinct mechanisms.

Anxiety, a non-motor symptom of PD that also reflects dopaminergic dysfunction, has been proposed as a factor associated with LID. For instance, Dias et al., in a cohort of 348 participants from the Parkinson’s Progression Markers Initiative (PPMI), reported that higher levels of trait anxiety preceded dyskinesia onset, as measured using the State-Trait Anxiety Inventory (STAI) [28]. Similarly, Eusebi et al., also utilizing data from PPMI of 398 participants, found that higher trait anxiety, measured by the STAI-Trait subscore, was associated with increased risk of dyskinesia [7].

In our study, although anxiety was self-reported by participants and not assessed using standardized instruments, we were still able to replicate its association with LID, even after adjusting for other variables in the multivariable model. This finding suggests that anxiety may play a relevant role in the development of dyskinesia and should be considered during LID risk assessment.

While some studies in non-Latino populations have reported a higher risk of LID among females, we did not observe this association (Supplementary Table 2; Supplementary Figure 4). Our findings align with previous studies conducted in Latin American countries, specifically Mexico and Brazil [9,17,18] where no association between LID and sex was identified. This suggests that sex-related differences in LID risk may vary across populations, probably due to genetic, environmental, or healthcare system-related factors.

The association between dopamine agonist use and dyskinesia has been widely debated. More recent evidence suggests that when dopamine agonists are used concurrently with levodopa, they may exacerbate existing dyskinesias. However, this effect may not stem from dopamine agonists alone, but rather from the concomitant use of levodopa. Dopamine agonists do not extend their effect into or beyond levodopa’s optimal therapeutic window and, on their own, are less likely to trigger LID [6]. Our findings align with this, suggesting that the total number of years on dopamine agonists, is not associated with LID after adjusting for LEDD and levodopa treatment duration.

A similar debate has been raised regarding the relevance of levodopa treatment duration and LID, with some studies stating that longer levodopa therapies increase the risk of developing LID [14], while others discarding this association [29]. In our study, a longer duration of levodopa therapy appeared to be significantly associated with LID, independently and when adjusting for other covariates such as disease duration and LEDD (Table 4).

However, given the cross-sectional nature of our study and the ongoing debate on these topics, these findings should be interpreted with caution.

Our study has several limitations. First, LID occurrence was recorded through a single self-reported or clinical observation rather than using a standardized assessment tool such as UPDRS Part IV. This cross-sectional approach may introduce recall bias and limits the ability to capture additional characteristics such as dyskinesia severity, impact on quality of life, time to onset (time-to-LID), and specific subtypes (e.g., peak-dose dyskinesia, diphasic dyskinesia, or off-period dystonia). Second, the calculated LEDD may underestimate the actual dopaminergic load, as data on medications other than levodopa and dopamine agonists were unavailable, as well as details on levodopa formulations (e.g., immediate-vs. extended-release). Additionally, information on other LID-targeted therapies, such as DBS, amantadine, or clozapine, was not available, and the use of these therapies could significantly influence the observed prevalence of LID in a cross-sectional analysis.

In conclusion, our findings represent the largest study to date on the epidemiology of LID in Latin American and the Caribbean countries. We demonstrate that several previously known dyskinesia-associated factors remain relevant in Latino populations, while also identifying potential new contributors, such as the clinical progression pattern of PD as defined in our cohort (fast, slow, or typical progressors). The absence of some expected associations, such as female sex, emphasizes the importance of population-specific research on LID and highlights the need for regionally tailored strategies for its prevention and management. Future studies incorporating more detailed clinical characterizations of dyskinesias, along with environmental, expositional, and genetic data, are needed to better understand the underlying mechanisms of LID.

## Data Availability

All data produced in the present study are available upon reasonable request to the authors

## Acknowledgments

This work was supported by the American Parkinson Disease Association (APDA, 1282087) [H.M.C-S., M.I-M., I.F.M.]; the Michael J. Fox Foundation (MJFF-026283) [E.W., I.F.M.]; the Parkinson’s Foundation (PDGENE-1333334) [M.I-M., I.F.M.]; the Alzheimer’s Disease Sequencing Project (ADSP, 5U01AG076482-03) [E.W., M.I-M.]; the Veterans Affairs Puget Sound Healthcare System (5I01ABX005978-2) [T.P.L., I.F.M.]; the National Council for Scientific and Technological Development (CNPq, Brazil) [P.B-N., B.L.S-L., A.F.S.S.]; FAP UNIFESP [V.B.]; the National Institutes of Health (NIH, R01NS112499-01A1) [T.P.L., M.I-M., I.F.M.]; Universidad Nacional de Tucumán (PIUNT-D711) [C.L.A.]; the Programa de Apoyo a Proyectos de Investigación e Innovación Tecnológica-Universidad Nacional Autónoma de México (IN208622) [S.A.]; and the Secretaría de Ciencia, Humanidades, Tecnología e Innovación (Secihti, CF-2019/6390) [S.A.].

This project was also supported by the Global Parkinson’s Genetics Program (GP2), which is funded by the Aligning Science Across Parkinson’s (ASAP) initiative and implemented by The Michael J. Fox Foundation for Parkinson’s Research (https://gp2.org). A complete list of GP2 members is available at https://gp2.org

The authors thank Luis Aguilar, Alejandro León, and Jair García of the Laboratorio Nacional de Visualización Científica Avanzada at the Universidad Nacional Autónoma de México, as well as Carina Uribe Díaz, Christian Molina-Aguilar, and Alejandra Castillo Carbajal, for their technical support. We are also grateful to Denise Paredes from Clinic Sanatorio de la Trinidad. We acknowledge the DNA-Neurogenetics Bank of the Instituto Nacional de Ciencias Neurológicas (INCN) in Peru, the Biobank of Tissue and Fluids of the University of Chile, and Hospital Nacional San Rafael and Hospital Nacional Rosales in El Salvador, for their valuable support and collaboration in this study. Additional thanks to the Centro de Investigaciones Clínicas (CIC) from Fundación Valle del Lili, Colombia. Finally, we are deeply grateful to all participants in LARGE-PD, including people with Parkinson’s disease and their families, whose involvement made this work possible.

## LARGE-PD Consortium

Argentina: Emilia Mabel Gatto, Claudia Perandones, Martin Radrizani, Gustavo DaPrat, Natalia Gonzalez Rojas, Melisa Espindola, Martin Cesarini, Maria Valentina Muller, Carlos Matias López Razquin, Bibiana Pizarro, Lucia Wang, Clarisa Marchetti, Cesar Avila, Griselda Alvarado, Luciana Rojas-Vazquez, Juan Pablo Diaz-Rearte, Marcelo Kauffman, Sergio Rodriguez-Quiroga, Dolores Gonzalez-Moron, Pavel Alejandro Hernandez, Belen Ceballos, Florencia Echeverria, Gabriela Costa, Marcelo Merello, Federico Capparelli, Florencia Wainberg, Tomas Poklepovich, Florencia Mallou, Denise De Belder, Jose Luis Etcheverry, Nelida Garretto, Tomoko Arakaki

Bolivia: Erick Gonzalez, Enrique Wagner, Robin Rodriguez, Alexander Quecana Janco

Brazil: Bruno Lopes Santos-Lobato, Gracivane Lopes Eufraseo, Juliana Santos Duarte, Marcella Montenegro, Tatiane Souza, Camille Sena, Ândrea Ribeiro-dos-Santos, Pedro Braga-Neto, Deborah Rangel, Marconny Cavalcante, Mateus Balsells, Vitor Tumas, Angela Vieira Pimentel, Vanderci Borges, Carolina Candeias da Silva, Henrique Ballali Ferraz, Dayany Leonel Boone, Mariana Cavalcanti Costa, Egberto Reis Barbosa, Grace Letro, Artur F. S. Schuh, Carlos R. M. Rieder, Gabriela Magalhães Pereira, Deise Cristine Friedrich, Thalya Osmilda Alves de Carvalho, Isabella Fonseca Benati, Jullivan Käfer Pasin, Vitor Picanço Lima Gomes, Marcelo Somma Tessari, Ingrid Lorena da Silva Gomes, Juan Sebastián Sánchez León, Paula Saffie-Awad, Gabriel Alves Marconi, Manoella Guatimuzim Testa da Silva, Gabriel Rosa Vilela, Eduardo Drews Amorim, Rafael Sidônio Gibson Gomes, André Vitor Souza Fernandes, Raphael Breder

Colombia: Gonzalo Arboleda, Oscar Bernal Pacheco, Norma Tatiana Lopez Gonzalez, Humberto Arboleda, Carlos Eduardo Arboleda Bustos, Hebert Bernal Castro, Juan David Caicedo Narvaez, Kelly Bonilla Vargas, Jorge Orozco, Beatriz Munoz Ospina, Harold Londono, David Aguillon, Sonia Moreno, Omar Buritica, David Pineda, Marlene Jimenez-Del-Rio, Carlos Vélez-Pardo, Sarita Firstman

Chile: Pedro Chana-Cuevas, Ximena Pizarro Correa, Consuelo Moos, Natalia Rojas, Patricio Olguin, Alicia Colombo, Juan Cristobal Nuñez, Andres De la Cerda, María Francisca Canals, Gonzalo Farías, Valentina Bessa, Mérida Terán, Pen Cheng Zhongxomg, Paula Saffie, Eduardo Perez, Dominga Berrios, Elías Fernandez-Toledo, Marlene Valenzuela-Valenzuela, Mario Fuentealba Sandoval, Susana Pineda, Floria Pancetti, Maria Eugenia Contreras Pinto, Benjamin Soto Flores

Costa Rica: Gabriel Torrealba-Acosta, Tanya Lobo-Prada, Jaime Fornaguera-Trías, Álvaro Hernandez-Guillen, Roger Rodríguez

Dominican Republic: Rossy Cruz Vicioso, Ernestina Castro, Alpher Perez, Sergio Mosquera, Cesarina Torres, Janfreisy Carbonell Honduras: Reyna M. Durón, Alex Medina, Heike Hesse, Evelin Alvarez Herrera, Eduardo P. Murillo, Glenda Oliva

El Salvador: Susana Pena, Tatiana Ascencio, Oscar Peña Rodas

Ecuador: Faryd Llerena Toro, Michael Castelo, Carlos Rodriguez

Mexico: Mayela Rodríguez-Violante, Ana Jimena Hernández-Medrano, Amin Cervantes-Arriaga, Daniel Martinez Ramirez, Sarael Alcauter, Alejandra Medina-Rivera, Alejandra E. Ruiz-Contreras, Miguel E. Rentería, Alejandra Lázaro-Figueroa, Juan Manuel Esquivias-Farias, Andrés Morales-de-Arcia, Alejandra Zayas-Del Moral, Damaris Vazquez-Guevara, Yamil Matuk-Pérez, Carlos Manuel Guerra-Galicia, Ildefonso Rodriguez-Leyva, Karla Salinas-Barboza, Eugenia Morelos-Figaredo, Omar Cardenas-Saenz, Nadia A Gandarilla-Martínez, Sara Isais-Millán, Teresa Pérez-Torres, Domingo Martinez, Ingrid Estada-Bellmann, Roberto Trejo-Ayala, Carlos Alberto Ponce-Fernández, Dante Oropeza

Peru: Mario Cornejo-Olivas, Julia Rios Pinto, Angel Medina, Ivan Cornejo-Herrera, Koni Mejia-Rojas, Cintia Armas Puente, Edward Ochoa-Valle, Marcela Alvarado Morales, Elison Sarapura Castro, Andrea Rivera-Valdivia, Maryenela Illanes Manrique, Carla Manrique Enciso, Victoria Marca Ysabel, Olimpio Ortega Dávila, Freddy Requejo-Navarro, Alid Manrique Palomino, Gabriela Gushiken Oshiro, Laura Zelada Rios

Uruguay: Elena Dieguez, Victor Raggio, Andres Lescano

USA-Puerto Rico: Angel Vinuela, Esther Colon

USA: Henry Mauricio Chaparro-Solano, Thiago P. Leal, Emily Waldo, Felipe Duarte-Zambrano, Miguel Inca-Martinez, Janvi Ramchandra, Kamilah Stark, Daniel Teixeira-dos-Santos, Emily

Leininger, Nicolas Gutierrez, Valerie Rico, Paula Reyes-Pérez, Maria Rivera Paz, Ignacio F. Mata, Karen Nuytemans, Anisley Martinez, Liena Infante

## REFERENCES

[1] Armstrong MJ, Okun MS. Diagnosis and Treatment of Parkinson Disease: A Review. JAMA 2020;323:548–60. 10.1001/jama.2019.22360.

[2] Aradi SD, Hauser RA. Medical Management and Prevention of Motor Complications in Parkinson’s Disease. Neurotherapeutics 2020;17:1339–65. 10.1007/s13311-020-00889-4.

[3] Uitti RJ, Ahlskog JE, Maraganore DM, Muenter MD, Atkinson EJ, Cha RH, et al. Levodopa therapy and survival in idiopathic Parkinson’s disease: Olmsted County project. Neurology 1993;43:1918–26. 10.1212/wnl.43.10.1918.

[4] Tanner CM, Ostrem JL. Parkinson’s Disease. N Engl J Med 2024;391:442–52. 10.1056/NEJMra2401857.

[5] di Biase L, Pecoraro PM, Carbone SP, Caminiti ML, Di Lazzaro V. Levodopa-Induced Dyskinesias in Parkinson’s Disease: An Overview on Pathophysiology, Clinical Manifestations, Therapy Management Strategies and Future Directions. J Clin Med 2023;12:4427. 10.3390/jcm12134427.

[6] Espay AJ, Morgante F, Merola A, Fasano A, Marsili L, Fox SH, et al. Levodopa-induced dyskinesia in Parkinson disease: Current and evolving concepts. Ann Neurol 2018;84:797–811. 10.1002/ana.25364.

[7] Eusebi P, Romoli M, Paoletti FP, Tambasco N, Calabresi P, Parnetti L. Risk factors of levodopa-induced dyskinesia in Parkinson’s disease: results from the PPMI cohort. NPJ Parkinsons Dis 2018;4:33. 10.1038/s41531-018-0069-x.

[8] Cenci MA, Riggare S, Pahwa R, Eidelberg D, Hauser RA. Dyskinesia Matters. Movement Disorders 2020;35:392–6. 10.1002/mds.27959.

[9] Tumas V, Brito MMCM, Borges V, Ferraz HB, Zabetian CP, Mata IF, et al. Levodopa-induced dyskinesia is still a major clinical problem in Brazilian movement disorder clinics. Arq Neuropsiquiatr 2025;83:1–5. 10.1055/s-0045-1806922.

[10] Turcano P, Mielke MM, Bower JH, Parisi JE, Cutsforth-Gregory JK, Ahlskog JE, et al. Levodopa-induced dyskinesia in Parkinson disease: A population-based cohort study. Neurology 2018;91:e2238–43. 10.1212/WNL.0000000000006643.

[11] Scott NW, Macleod AD, Counsell CE. Motor complications in an incident Parkinson’s disease cohort. Eur J Neurol 2016;23:304–12. 10.1111/ene.12751.

[12] Martinez-Carrasco A, Real R, Lawton M, Iwaki H, Tan MMX, Wu L, et al. Genetic meta-analysis of levodopa induced dyskinesia in Parkinson’s disease. NPJ Parkinsons Dis 2023;9:128. 10.1038/s41531-023-00573-2.

[13] Gandhi SE, Zerenner T, Nodehi A, Lawton MA, Marshall V, Al‐Hajraf F, et al. Motor Complications in Parkinson’s Disease: Results from 3343 Patients Followed for up to 12 Years. Mov Disord Clin Pract 2024;11:686–97. 10.1002/mdc3.14044.

[14] Santos-García D, de Deus T, Cores C, Feal Painceiras MJ, íñiguez Alvarado MC, Samaniego LB, et al. Levodopa-Induced Dyskinesias are Frequent and Impact Quality of Life in Parkinson’s Disease: A 5-Year Follow-Up Study. Mov Disord Clin Pract 2024;11:830–49. 10.1002/mdc3.14056.

[15] Chang HJ, Jang M, Woo KA, Shin JH, Kim H-J, Jeon B. The prevalence of non-troublesome dyskinesia in Parkinson’s disease. Parkinsonism Relat Disord 2024;123:106951. 10.1016/j.parkreldis.2024.106951.

[16] Loo RTJ, Tsurkalenko O, Klucken J, Mangone G, Khoury F, Vidailhet M, et al. Levodopa-induced dyskinesia in Parkinson’s disease: Insights from cross-cohort prognostic analysis using machine learning. Parkinsonism Relat Disord 2024;126:107054. 10.1016/j.parkreldis.2024.107054.

[17] Santos-Lobato BL, Schumacher-Schuh AF, Rieder CRM, Hutz MH, Borges V, Ferraz HB, et al. Diagnostic prediction model for levodopa-induced dyskinesia in Parkinson’s disease. Arq Neuropsiquiatr 2020;78:206–16. 10.1590/0004-282X20190191.

[18] Cervantes-Arriaga A, Rodríguez-Violante M, Salmerón-Mercado M, Calleja-Castillo J, Corona T, Yescas P, et al. [Incidence and determinants of levodopa-induced dyskinesia in a retrospective cohort of Mexican patients with Parkinson’s disease]. Rev Invest Clin 2012;64:220–6.

[19] Zabetian CP, Mata IF, Latin American Research Consortium on the Genetics of PD (LARGE-PD). LARGE-PD: Examining the genetics of Parkinson’s disease in Latin America. Mov Disord 2017;32:1330–1. 10.1002/mds.27081.

[20] Zhang Z. Multiple imputation with multivariate imputation by chained equation (MICE) package. Ann Transl Med 2016;4:30. 10.3978/j.issn.2305-5839.2015.12.63.

[21] Mehanna R, Smilowska K, Fleisher J, Post B, Hatano T, Pimentel Piemonte ME, et al. Age Cutoff for Early-Onset Parkinson’s Disease: Recommendations from the International Parkinson and Movement Disorder Society Task Force on Early Onset Parkinson’s Disease. Movement Disorders Clinical Practice 2022;9:869–78. 10.1002/mdc3.13523.

[22] Zhao YJ, Wee HL, Chan Y-H, Seah SH, Au WL, Lau PN, et al. Progression of Parkinson’s disease as evaluated by Hoehn and Yahr stage transition times. Mov Disord 2010;25:710–6. 10.1002/mds.22875.

[23] Jost ST, Kaldenbach M-A, Antonini A, Martinez-Martin P, Timmermann L, Odin P, et al. Levodopa Dose Equivalency in Parkinson’s Disease: Updated Systematic Review and Proposals. Mov Disord 2023;38:1236–52. 10.1002/mds.29410.

[24] Sosero YL, Bandres-Ciga S, Ferwerda B, Tocino MTP, Belloso DR, Gómez-Garre P, et al. Dopamine Pathway and Parkinson’s Risk Variants Are Associated with Levodopa-Induced Dyskinesia. Mov Disord 2024;39:1773–83. 10.1002/mds.29960.

[25] Schlickmann TH, Tessari MS, Borelli WV, Marconi GA, Pereira GM, Zimmer E, et al. Prevalence, distribution and future projections of Parkinson disease in Brazil: insights from the ELSI-Brazil cohort study. The Lancet Regional Health – Americas 2025;44. 10.1016/j.lana.2025.101046.

[26] Riederer P, Strobel S, Nagatsu T, Watanabe H, Chen X, Löschmann P-A, et al. Levodopa treatment: impacts and mechanisms throughout Parkinson’s disease progression. J Neural Transm (Vienna) 2025;132:743–79. 10.1007/s00702-025-02893-4.

[27] Bovenzi R, Conti M, Degoli GR, Cerroni R, Simonetta C, Liguori C, et al. Shaping the course of early-onset Parkinson’s disease: insights from a longitudinal cohort. Neurol Sci 2023;44:3151–9. 10.1007/s10072-023-06826-5.

[28] Dias CMV, Leal DAB, Brys I. Levodopa-induced dyskinesia is preceded by increased levels of anxiety and motor impairment in Parkinson’s disease patients. International Journal of Neuroscience 2023;133:1319–25. 10.1080/00207454.2022.2079501.

[29] Giannakis A, Chondrogiorgi M, Tsironis C, Tatsioni A, Konitsiotis S. Levodopa-induced dyskinesia in Parkinson’s disease: still no proof? A meta-analysis. J Neural Transm (Vienna) 2018;125:1341–9. 10.1007/s00702-018-1841-0.

