## Supplementary Material for "EPIDEMIOLOGY OF LEVODOPA-INDUCED DYSKINESIA: PREVALENCE AND ASSOCIATED CLINICAL FACTORS IN LATIN AMERICA"

**SUPPLEMENTARY TABLES**

| **Supplementary table 1. LID prevalence by disease duration and age at onset group** | | | |
| --- | --- | --- | --- |
| **Disease duration group** | **Age at onset <50** | **Age at onset ≥50** | ***p*-value** |
| 0-5 years | 16.8% (N=244) | 7% (N=1112) | 1.85E-06 |
| 6-10 years | 38.7% (N=344) | 19.4% (N=823) | 8.48E-12 |
| 11-15 years | 52.8% (N=252) | 30.7% (N=423) | 2.15E-08 |
| 16-20 years | 61.8% (N=152) | 40% (N=125) | 4.65E-04 |
| > 20 years | 56.2% (N=176) | 50% (N=44) | 5.65E-01 |

| **Supplementary table 2. LID prevalence by disease duration and sex** | | | |
| --- | --- | --- | --- |
| **Disease duration group** | **Female** | **Male** | ***p*-value** |
| 0-5 years | 9.2% (N=590) | 8.5% (N=766) | 7.39E-01 |
| 6-10 years | 24.1% (N=465) | 25.8% (N=702) | 5.58E-01 |
| 11-15 years | 35.6% (N=253) | 41% (N=422) | 1.88E-01 |
| 16-20 years | 58% (N=119) | 47.5% (N=158) | 1.07E-01 |
| > 20 years | 61.9% (N=97) | 49.6% (N=123) | 9.32E-02 |

**SUPPLEMENTARY FIGURES**

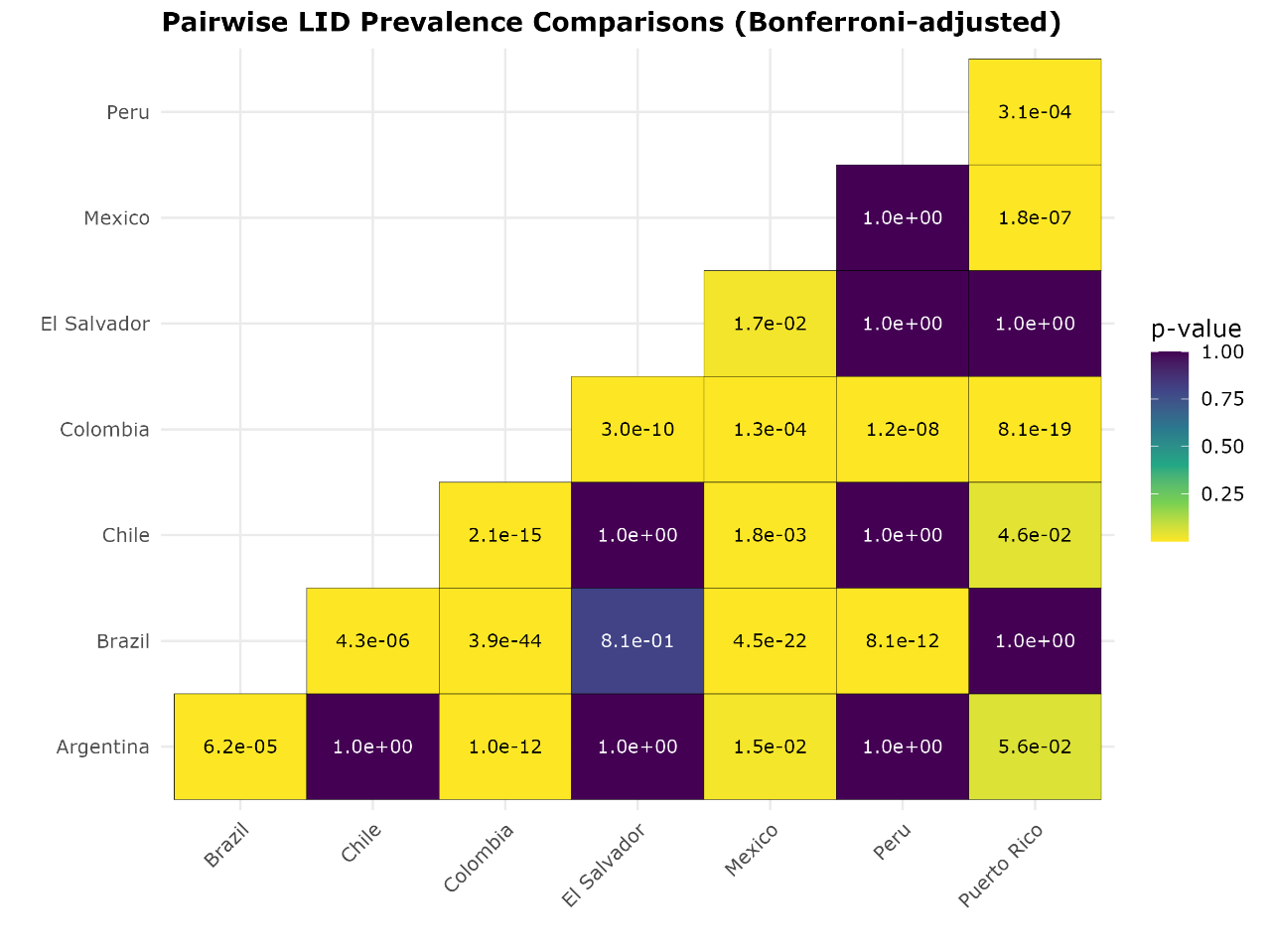

**Supplementary Figure 1. Bonferroni-adjusted pairwise comparisons of Levodopa-induced dyskinesia (LID) prevalence between countries.** Each tile represents the *p*-value from a pairwise comparison, with color indicating the magnitude of the *p*-value (yellow = lower, purple = higher). *P*-values are displayed within tiles; comparisons with *p* < 0.05 after adjustment are considered statistically significant.

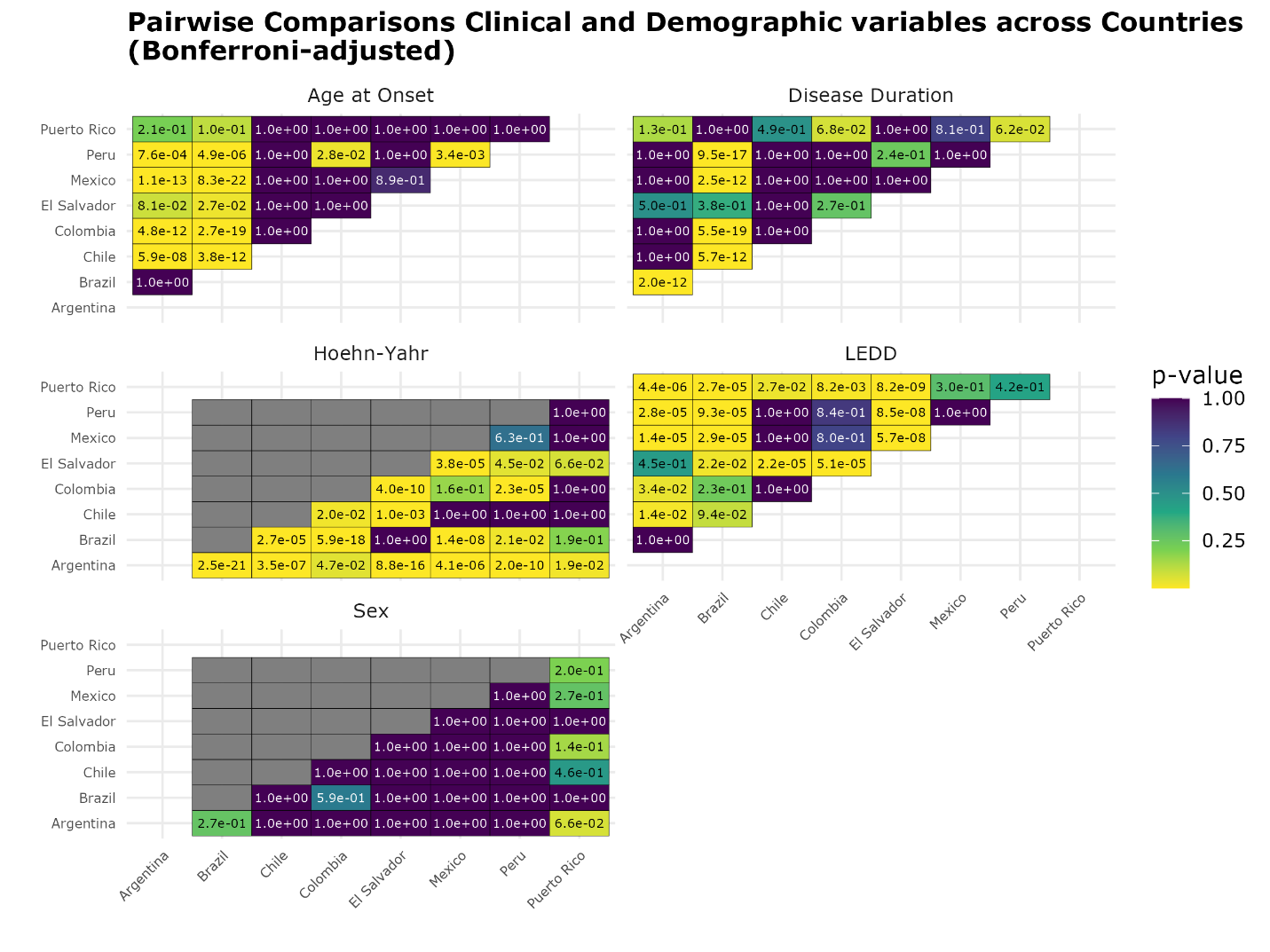

**Supplementary Figure 2. Bonferroni-adjusted pairwise comparisons of clinical and demographic variables between countries.** Each panel corresponds to a different variable assessed in the analysis. Each tile displays the adjusted *p*-value for a specific variable comparison between two countries, with color indicating the magnitude of the *p*-value (yellow = lower, purple = higher). *P*-values are shown within the tiles; comparisons with *p* < 0.05 after adjustment are considered statistically significant.

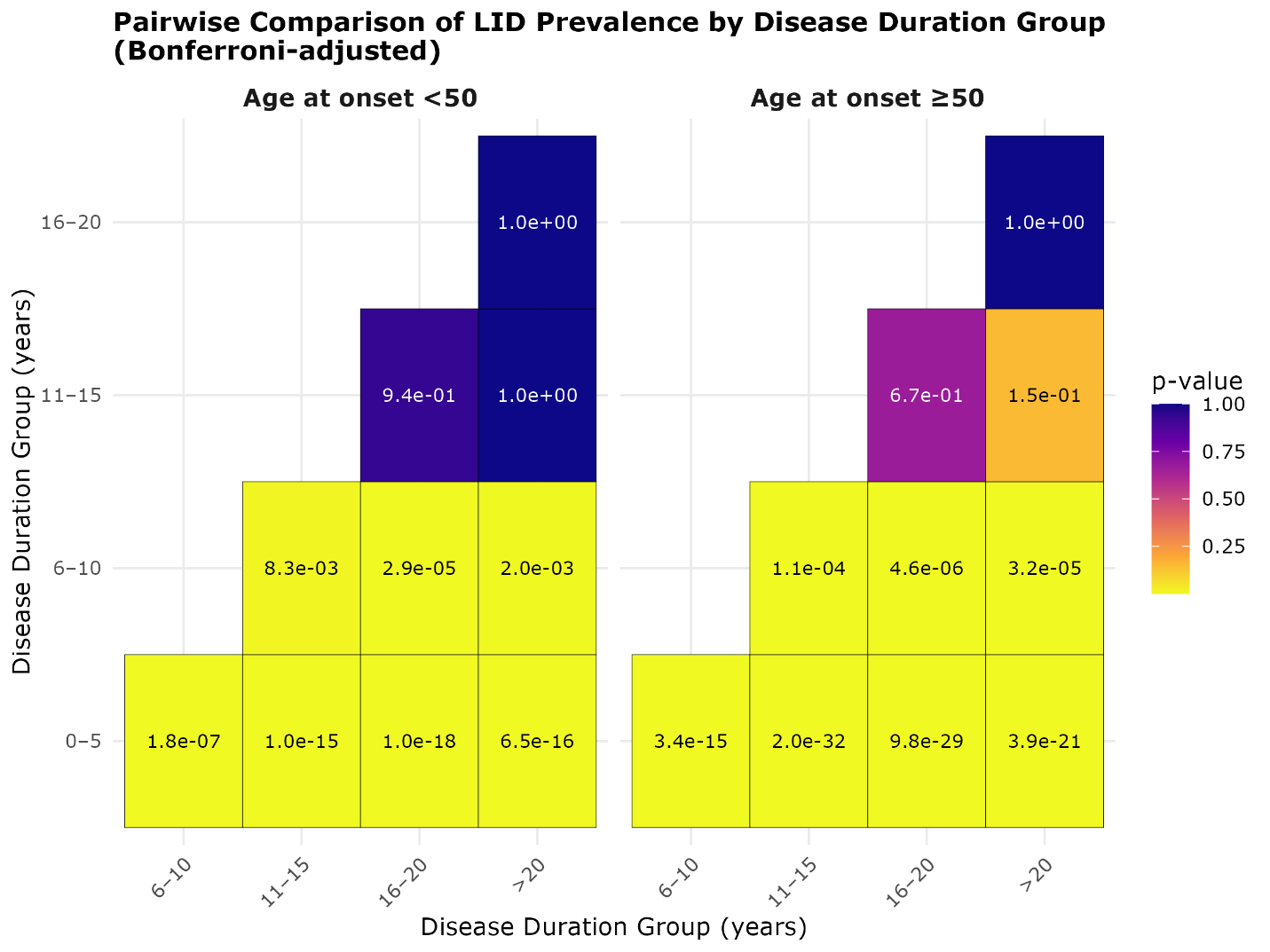

**Supplementary Figure 3. Bonferroni-adjusted pairwise comparisons Levodopa-induced dyskinesia (LID) prevalence across disease duration groups, stratified by age at onset**. Each tile represents a comparison between two disease duration groups within the same age group, with *p*-values shown inside the tiles and color indicating their magnitude (yellow = lower, purple = higher). Comparisons with *p* < 0.05 after adjustment are considered statistically significant. Panels represent different age-at-onset categories.

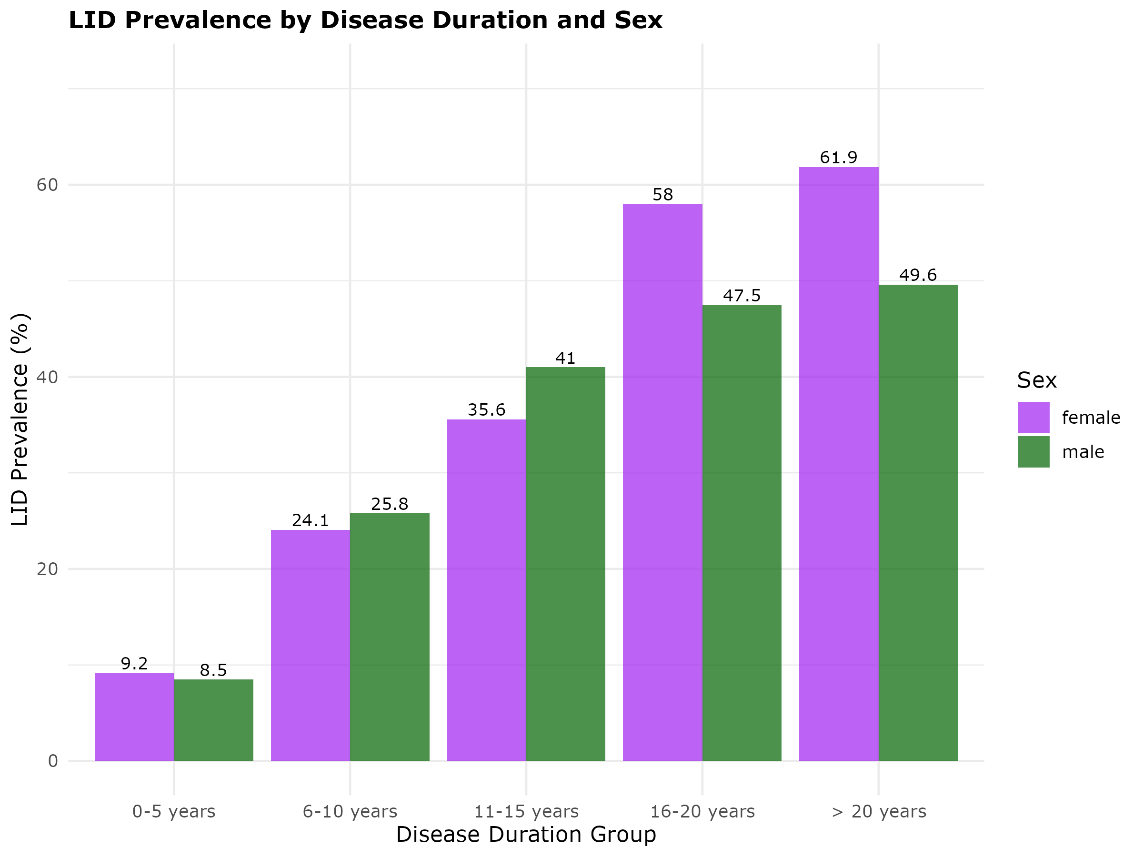

**Supplementary Figure 4. Levodopa-induced dyskinesia (LID) prevalence by disease duration and sex.** Prevalence of LID (Y-axis) across different Parkinson’s disease duration groups (X-axis), stratified by sex. Purple bars, female sex, green bars, male sex.
